# Prevalence and time trend of congenital heart defects: a registry-based study in Iran

**DOI:** 10.1101/2021.12.16.21267826

**Authors:** Hossein Molapour, Saeed Dastgiri, Elham Davtalab Esmaeili

**Affiliations:** Medical student, School of Medicine, Tabriz University of Medical Science, Tabriz, Iran; Tabriz Registry of Congenital Anomalies (TRoCA), and Tabriz Health Services Management Research Center, School of Medicine, Tabriz University of Medical Sciences, Tabriz, Iran; Road Traffic Injury Research Center, Tabriz University of Medical Sciences, Tabriz, Iran

**Keywords:** Congenital heart defects, Prevalence, Congenital heart anomalies, Epidemiology

## Abstract

**Introduction:** Congenital Heart Defects (CHD) are one of the most common types of congenital anomalies affecting nearly one percent of births every year. The aim of this study was to report the prevalence and time trend of CHD in the northwest of Iran between 2000 and 2019.

**Methods:** Since 2000, infants born with birth defects have been registered in Tabriz Registry of Congenital Anomalies (TRoCA). For this study, the information and data of newborns with CHD (n=1084) were collected using the TRoCA registry system.

**Results:** Over the two decades, the prevalence rate of CHD in the northwest of Iran was 3.7 per 1000 live births (95 percent CI: 34.9 to 39.4). The prevalence rate of CHD during the first decade (2000-09) and the second (2010-19) were estimated 2.8 (95 percent CI: 2.5 to 2.9) and 5.9 (95 percent CI: 5.4 to 6.4) per 1000 live births, respectively.

**Conclusion:** The prevalence rate of CHD in the TRoCA registry area showed an increasing trend indicating methodological improvement in the facilities and diagnosis techniques. It would therefore seem essential to concentrate on the primary prevention activities to reduce the burden of these defects.

## Introduction

Congenital Heart Defects (CHD) are common congenital defects accounting for one-third of major congenital malformations, and a leading cause for some comorbidities, mortality, and abortions.^1-3^ The prevalence of CHD was reported 5.2-10.7 (per 1,000 live births) from different countries. ^4, 5^ According to the Center for Disease Control and Prevention (CDC), CHD affects one percent of all births in the United States. ^6^ World Health Organization (WHO) reported the total prevalence of congenital anomalies is about 65 (per 1,000 live births) in the Eastern Mediterranean region from which 40 percent is related to CHD. In Iran, the prevalence of CHD is estimated from 7.93 to 17.51 (per 1,000 live births).^7^ Almost 34 percent of neonatal with a major CHD die within the first month of life if they didn’t access appropriate intervention.^8, 9^ It is estimated that 60 percent of CHD patients could be treated if early diagnosis and adequate health facilities are available. ^10^

Determining the prevalence of CHD trend over time may provide an insight for policymakers to develop health prevention programms. This registry-based study aimed to investigate the prevalence and time trend of CHD in the northwest of Iran over two decades between 2000 and 2019.

## Materials and methods

Since 2000, all infants born with congenital anomalies including CHD have been registered in the Tabriz Registry of Congenital Anomalies (TRoCA) in the northwest of Iran. The registry has been accepted in the International Clearinghouse for Birth Defects Surveillance and Research (ICBDSR), as a member of countries having an established registry for birth defects. It is also a world affiliate member of the European network of registries for the epidemiologic surveillance of congenital anomalies (EUROCAT).

TRoCA is a hospital-based registry which is located in the northwest of Iran, Tabriz city, covering all births and children in three university referral hospitals. Those hospitals provide obstetric and gynecological services in the population of TRoCA region. All newborns are routinely examined by a gynecologist, obstetrician or neonatologist at birth and hospital discharge. The examinations include assessment of general health, maturity and the existence of birth defects including CHD. Epidemiological information are also collected for every newborn and for mothers of all malformed infants. The majority of deliveries (70 percent) among general population plus high risk pregnancies are carried out in those public university hospitals. There are less than 2 percent of home delivery in the country.^11^

For the purpose of this study, the information for infants born with CHD including gender, birth weight, maternal age, consanguinity, ABO and Rh blood group, place of living and the presence of other congenital anomalies over two decades between 2000 and 2019 were gathered using TRoCA registry. Anomalies were coded using the International Classification of Diseases coding system (Version 10) and 95 percent confidence intervals were calculated to assess the statistical significance of the data. Statistical analysis was performed using the MS Excel and Graph Pad Prism software (version 6.0).

The Present study protocol was approved by the ethics committee of the Tabriz University of Medical Sciences (IR.TBZMED.REC.1399.625).

## Results

A total of 291569 live births were investigated by TRoCA registry from which 1084 cases have a primary diagnosis of CHD representing an overall prevalence rate of 3.7 per 1000 live births (95 percent CI: 3.4 to 3.9). Figure 1 shows the prevalence and the time trend of CHD in the area during the study period. The prevalence rate of CHD during the first decade (2000-09) and the second (2010-19) were estimated 2.8 (95 percent CI: 2.5 to 2.9) and 5.9 (95 percent CI: 5.4 to 6.4) per 1000 live births, respectively. The highest prevalence rate estimated for 2010 (10.4 per 1000; 95 percent CI: 9.1 to 11.7), and lowest prevalence rate belonged to the year 2001 (0.7 per 1000; 95 percent CI: 0.3 to 1.1).

**Figure 1.**
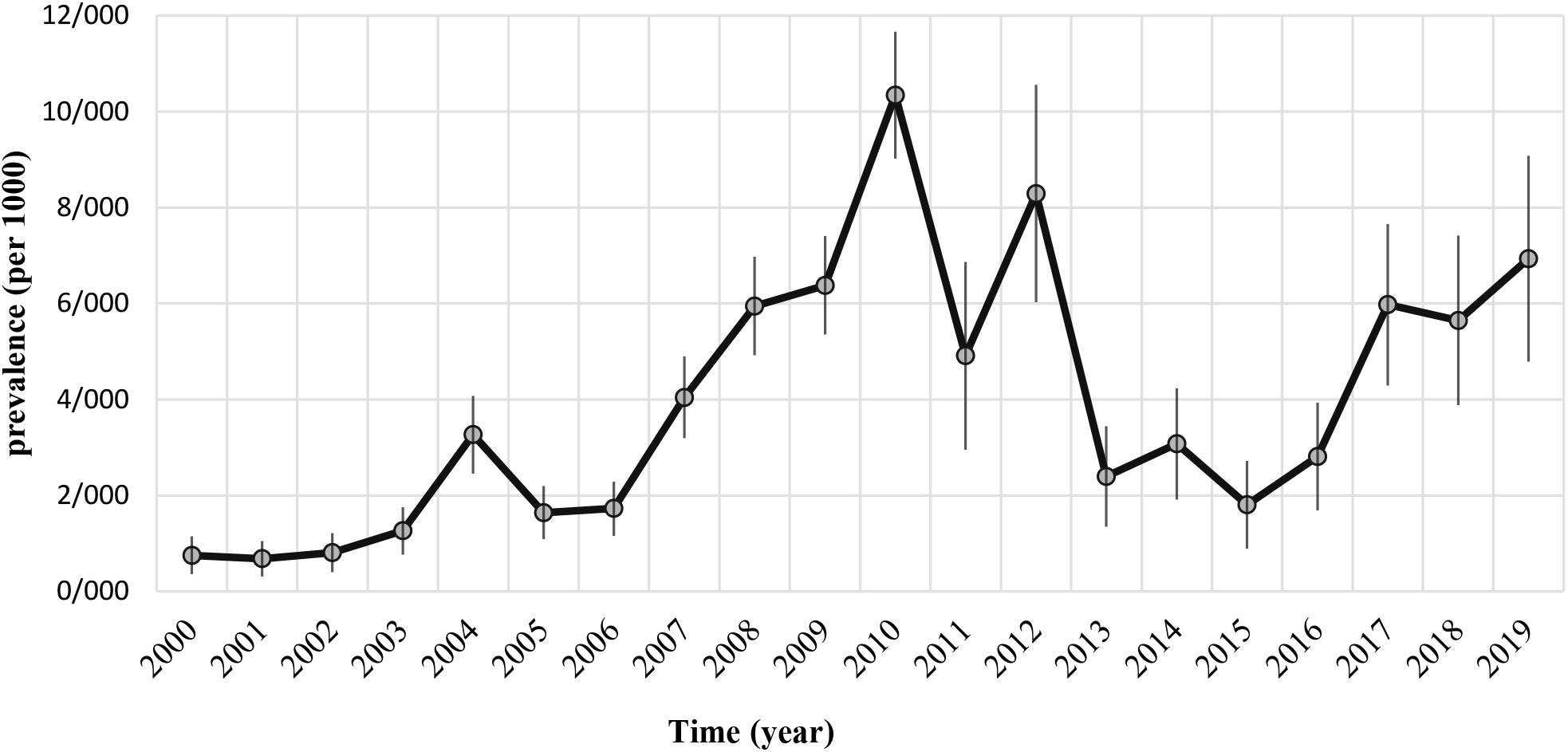
The prevalence (95 percent CI) and the time trend of congenital heart defects in the northwest of Iran (2000-19)

Occurrence of CHD was more common in boys than girls, representing a sex ratio (M/F) of 1.35. Cases who had more defects in addition to CHD was reported 181 (18.6 percent). The majority (72.7 percent) of parents of cases had no consanguineous marriage compared to 15.1 and 12.2 percent for the first and second cousin marriage among parents of study subjects. Type A+ and O+ blood groups were more common. Most infants with CHD (67.4 percent) had birth weight greater than 2500 grams, and most (92.1 percent) born from mother between age 18 to 35 (Table 1).

**Table 1.**
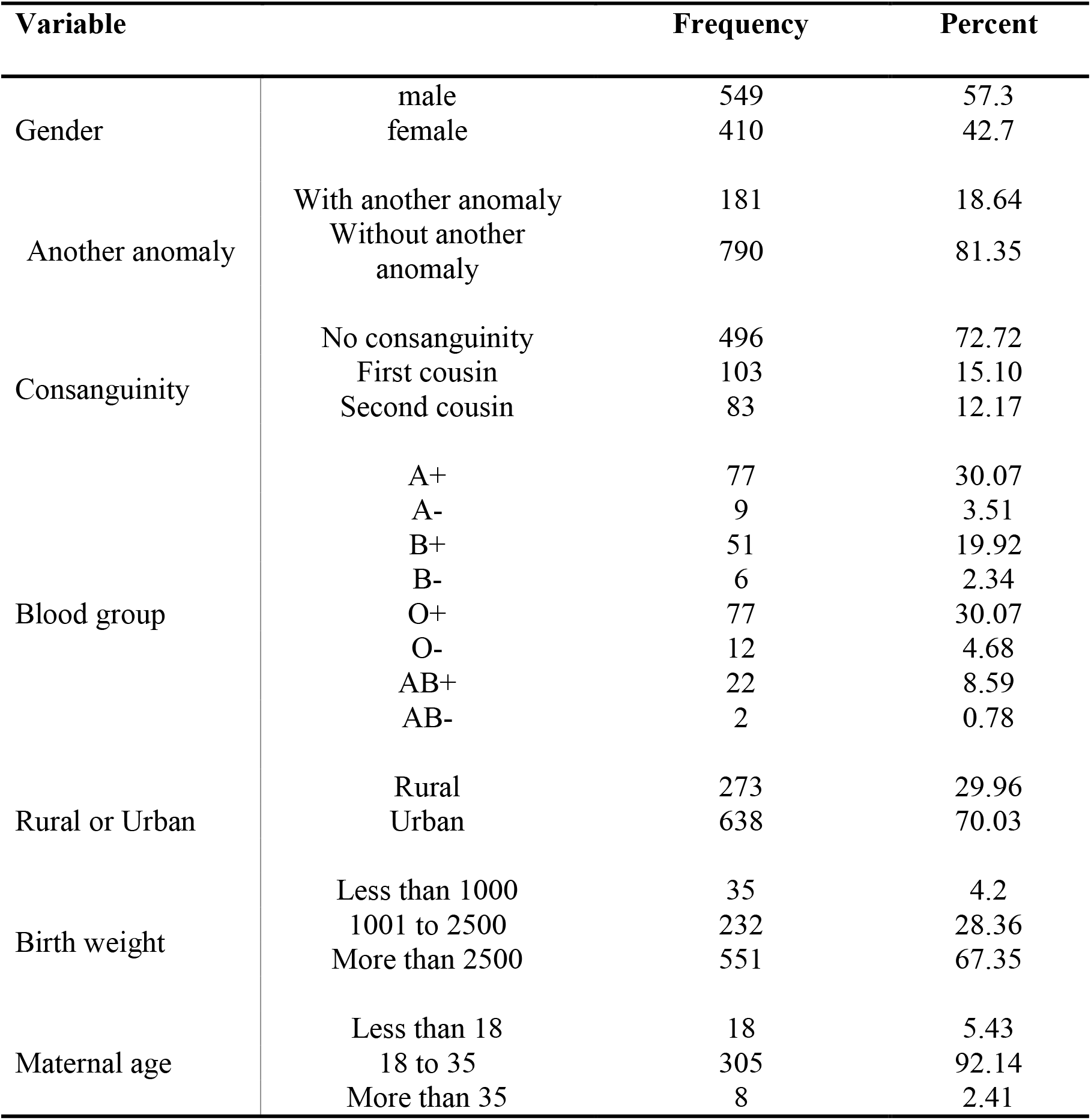
characteristics of infant with CHD at born during 2000 to 2019 in the northwest of Iran

## Discussion

According to our results, the prevalence of CHD in the northwest of Iran had an increasing trend (2.14 folds) over the study years.

Some studies found significant geographical differences in the prevalence of CHD worldwide.^2^ The prevalence rate of CHD in China, Kuwait, India and Egypt was 5.0, 8.9, 6.6, 13.0 and 1.0 (per 1000 live births), respectively.^12-15^ According to the study of Dolk et al the prevalence rate of CHD in Belgium, Croatia, Denmark, Germany, Ireland, Italy, the Netherlands, Norway, Poland, Spain, Sweden, Ukraine and the United Kingdom was 15.3, 5.2, 9.0, 11.6, 6.0, 6.8, 6.1, 10.2, 18.1, 5.5, 13.4, 7.7 and 6.8 (per 1000 live births), respectively.^16^ Most of this CHD prevalence rates is close to the findings of our study mainly in the second decade of the study years showing probably the diagnostic improvement for CHD in the region. During the past years, several studies in different parts of Iran have also investigated the prevalence of CHD with various figures from 16.2, 7.3, 13 and 4.2 per 1000 live births in Kashan, Tehran, Ahvaz and Khorramabad, respectively. ^17-19^

Variation of the CHD rates is mainly depending on the ability to detect and diagnosis of the defects in maternity settings in various regions.^21^ For instance, according to the study of Dastgiri et al (2011), almost 60 percent of children with congenital cardiac anomalies are not identified at birth in Iran.^21^ Therefore, the higher rate of CHD does not necessarily indicate a true increase in the incidence of CHD, and could rather be due to the improvements in the diagnostic ability in the children and maternal facilities.

Although some studies concluded that type A blood group could reduce the risk of isolated-CHD, type A blood group was more common than others in the present study.^22^ These findings need further investigations to confirm. The majority of the cases of CHD in our study belonged to urban areas indicating that higher education, economic level, life style and healthcare facilities might have a role in early detection of cardiac defects.

## Strengths, limitations and recommendations

Our study has some strengths including the use of registry data in a large population in the northwest of Iran, and the large sample (n=1084) used to estimate the prevalence and time trend of CHD in Iranian population. This study should however be considered in the light of some certain limitations. First, although there might be some suspected heterogeneous etiologies, we were unable to analyse the CHD subtypes separately. This may be the matter of another research in the future. Second, because of inability to fully adjust for various confounders, the increasing trend of the occurrence of CHD in our population of study could be attributed to some other factors including parental age, education and smoking, ethnicity, and some environmental exposures. It would also be recommended to investigate the role of those contributing factors in details in the future studies.

## Conclusion

In accordance with reports from various countries, the prevalence rate of CHD in Iran showed an increasing trend indicating methodological improvement in the facilities and diagnosis techniques. It would therefore seem essential to concentrate on the primary prevention activities to reduce the burden of these defects in the region and at the national level.

## Data Availability

All data produced in the present work are contained in the manuscript

## Acknowledgment

We gratefully thank all the children and mothers in the region for their assistance in TRoCA programme over the past two decades. This study has been funded by the Tabriz University of Medical Sciences (IR.TBZMED.REC.1399.625).

